# Evolving Virulence? Decreasing COVID-19 Complications among Massachusetts Healthcare Workers: A Cohort Study

**DOI:** 10.1101/2020.08.17.20176636

**Authors:** Fan-Yun Lan, Robert Filler, Soni Mathew, Eirini Iliaki, Rebecca Osgood, Lou Ann Bruno-Murtha, Stefanos N. Kales

## Abstract

The hypothesis of attenuating SARS-CoV-2 virulence has been raised. We examined the temporal distribution of COVID-19 complications (ER visits, hospitalization, intubation/code, and/or death) among healthcare workers in one system that applied uniform screening criteria throughout the research period, and found the complication rate significantly decreased after April 15, 2020.

---

There is limited research investigating possible changes in SARS-CoV-2 virulence. Existing evidence shows a decreasing case-fatality rate in the United States*(1)*. However, in relation to virulence, this statistic may be biased by improved clinical acumen as well as increased testing detecting more asymptomatic and paucisymptomatic cases. An Italian study has demonstrated lower nasopharyngeal viral load in the later phase of the pandemic *(2)*. In addition, attenuated SARS-CoV-2 variants have been reported *(3,4)*. Therefore, we investigated the temporal distribution of COVID-19 complications among healthcare workers (HCWs) to determine if the clinical case severity decreased over time.

This retrospective cohort study examined adult HCWs from a Massachusetts community healthcare system using a uniform COVID-19 protocol to screen and test employees since the initial outbreak in March 2020. Inclusion criteria were: 1) COVID-19 triage via the occupational health “hotline” and a documentation of a positive SARS-CoV-2 viral assay between March 9 and July 8, 2020, or 2) any untriaged employee with a complication due to COVID-19 subsequently confirmed by the system’s occupational health department. Since the initial outbreak, the occupational health “hotline” phone-interviewed HCWs with any concerns regarding a) travel; b) potential contact with a COVID-19-positive/suspect person; or c) possible viral symptoms, and referred HCWs for a nasopharyngeal SARS-CoV-2 RT-PCR assay when clinically indicated. HCWs tested outside the system sent in their PCR results for confirmation. All cases were contacted regularly and followed until July 31, 2020 for any complication, including: ER (Emergency Room) visit, hospitalization, intubation/code, and/or death. Detailed data acquisition, de-identification and ethical statement have been published previously *(5)*.

Because Massachusetts has used April 15^th^ as the date to measure improvements from the states’ pandemic peak (Figure) *(6)*, we defined the cases triaged up to April 15, 2020, as “early”, and those afterwards as “late”. We also included as early, any untriaged death occurring within 18 days of April 15, 2020 based on the median time from infection onset to death *(7,8)*. Sensitivity analysis was done by restricting the study population to only triaged COVID-19 cases.

**Figure.**
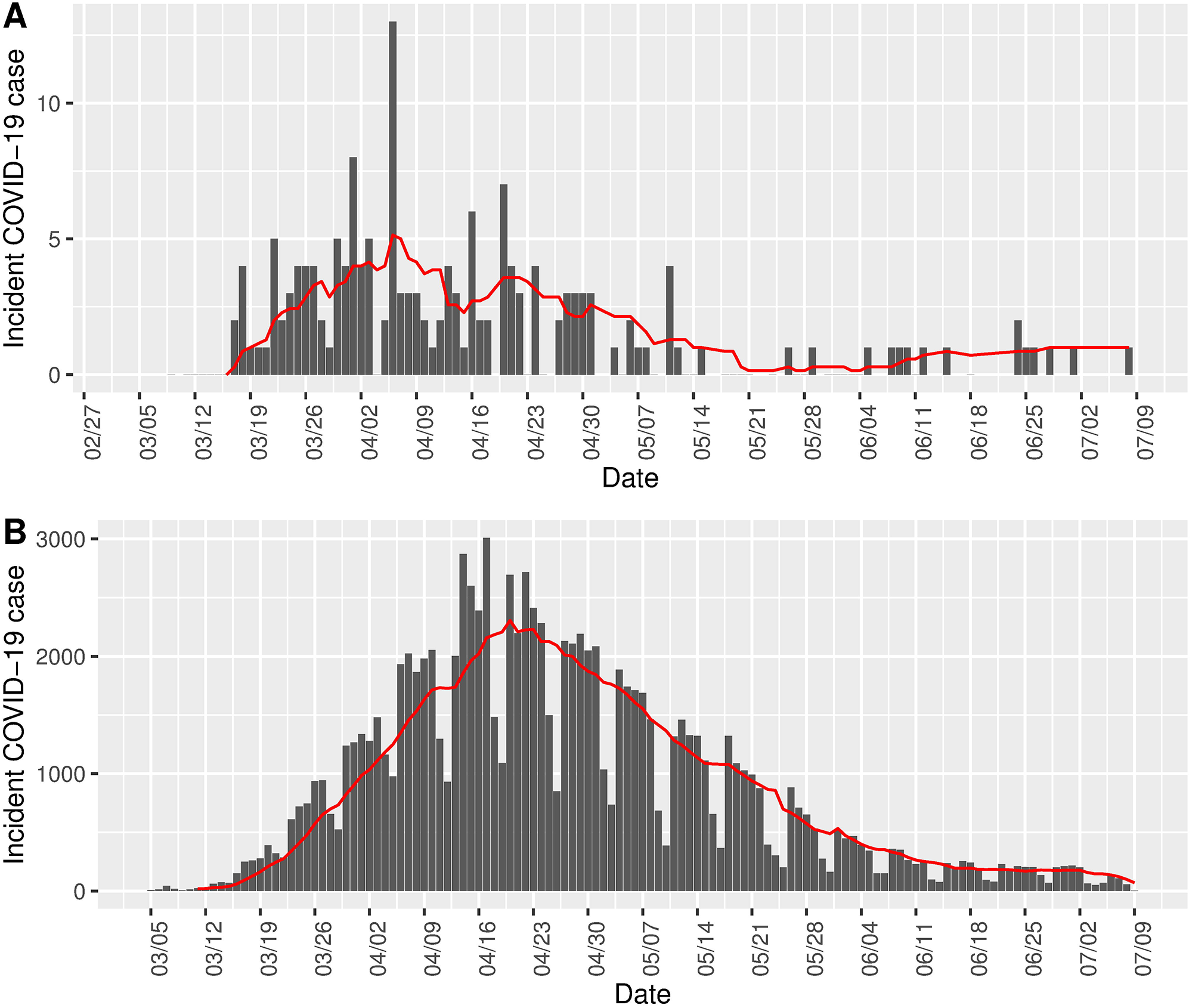
Temporal Distribution of Incident Confirmed COVID-19 Cases from March 5 to July 9, 2020 in. (A) the Study Population and (B) Massachusetts Statewide

Bars indicate the absolute number of daily cases, and the red line denotes the 7-day average of new cases.

We used the t-test or Wilcoxon rank sum test, as appropriate, to compare continuous variables, and chi-square test or the Fisher’s exact test, as appropriate, for categorical variables. Logistic regression models were built for multivariate adjustment. The analyses were performed using R software (version 3.6.3), two-sided and with a *P*-value < 0.05 considered statistically significant.

The temporal distribution of incident COVID-19 cases in the study population showed a pattern similar to that statewide*(6)* (Figure). A total of 166 eligible cases were identified in the study period: 98 early and 68 late. There were no significant differences between early and late cases regarding age, sex, race, residential community attack rates*(6)*, and number of presenting symptoms. Cough was more frequently reported in early cases (66% vs. 50%, *P* = 0.04), while headache was more frequently reported later (42% vs. 60%, *P* = 0.03). There were a total of 13 cases with any complications in the database, comprised of 12 early cases and one late case*(P* = 0.02). Ten cases experienced hospitalization or death (9 early vs. 1 late, *P*< 0.05). Three cases underwent intubation or a code (all early cases) and two of these HCWs died (Table). After adjusting for age, sex, and race, early cases had increased odds ratios (ORs) in any complication(s) (10.3, 95%CI: 1.7–199.9), and hospitalization/death (6.2, 95%CI: 1.0–122.5).

**Table.** Early and late COVID-19 cases’ clinical characteristics and complication rates.

|  | Early (thru 15 April)<br>(n=98) | Late (16 April–9 July)<br>(n=68) | P-value |
| --- | --- | --- | --- |
| Age (n=166) | 43.1 (12.9) | 41.5 (11.9) | 0.440 |
| Sex (n=159) |  |  | 0.944 |
| Female | 70 (71.4%) | 46 (67.6%) |  |
| Male | 25 (25.5%) | 18 (26.5%) |  |
| Race (n=156) |  |  | 0.517 |
| Non-Hispanic white | 38 (38.8%) | 18 (26.5%) |  |
| African American | 33 (33.7%) | 27 (39.7%) |  |
| Hispanic | 18 (18.4%) | 14 (20.6%) |  |
| Others | 5 (5.1%) | 3 (4.4%) |  |
| Cumulative community attack rate*<br>(as of June 3) (n=156) | 1723.7 (1214.2–3393.3) | 1802.8 (1634.5–2330.2) | 0.883† |
| Total number of symptoms<br>(n=165) | 4 (3–5) | 4 (2–5) | 0.529† |
| No symptom | 4 (4.1%) | 4 (5.9%) | 0.718‡ |
| Fever | 50 (51.0%) | 25 (36.8%) | 0.086 |
| Cough | 65 (66.3%) | 34 (50.0%) | **0.042** |
| Dyspnea | 13 (13.3%) | 12 (17.6%) | 0.598 |
| Myalgia | 48 (49.0%) | 34 (50.0%) | 0.999 |
| Malaise | 51 (52.0%) | 29 (42.6%) | 0.272 |
| Nasal symptoms | 33 (33.7%) | 28 (41.2%) | 0.439 |
| GI symptoms | 26 (26.5%) | 15 (22.1%) | 0.609 |
| Rash | 3 (3.1%) | 1 (1.5%) | 0.644‡ |
| Anosmia/Ageusia | 14 (14.3%) | 18 (26.5%) | 0.085 |
| Headache | 41 (41.8%) | 41 (60.3%) | **0.034** |
| Sore throat | 41 (41.8%) | 20 (29.4%) | 0.129 |
| Any complication(s) (n=166) | 12 (12.2%) | 1 (1.5%) | **0.016‡** |
| ER visit | 12 (12.2%) | 1 (1.5%) | **0.016‡** |
| Hospitalization/death§ | 9 (9.2%) | 1 (1.5%) | **0.049‡** |
| Intubation/coded§ | 3 (3.1%) | 0 | 0.270‡ |
| Death | 2 (2.0%) | 0 | 0.513‡ |

Mean (SD) for age. Median (Q1-Q3) for cumulative community attack rate and total number of symptoms. Count (%)
for other variables.
Nasal symptoms include runny, sneezing, congestion, and sinus symptoms. GI (Gastrointestinal) symptoms include
nausea/ vomiting/ diarrhea. ER, Emergency Room.
\*Community rates based on Massachusetts data for each healthcare worker's town of residence.
†Wilcoxon rank sum test
‡Fisher's exact test
§Patient coded and died in an Emergency Department.

In the sensitivity analysis, we excluded one COVID-19 death on April 29, who did not undergo triage and had an unknown date of symptom onset. The results remained significant regarding early versus late total complications: 11.3% (11/97) and 1.5% (1/68) *(P* = 0.02).

Our study provides epidemiologic support for the hypothesis of possible attenuated viral virulence *(2–4)* regarding clinical severity. We found cases infected later in the pandemic were less likely to sustain a complication. Later cases were also more likely to have headache, which may portend a more favorable course *(9)*. The current study has several strengths. First, we used uniform screening protocols and testing criteria for all employees throughout the study period, minimizing bias from younger and less symptomatic cases detected as testing became more widespread. In fact, we observed that the age, sociodemographic characteristics and presenting symptoms of early and late cases were in fact quite similar, and the results remained significant after adjusting for these potential confounders. Second, the similar temporal epidemic pattern between the HCWs and the general population implies possible generalizability to other working populations. Nonetheless, the present study had limited sample size and HCWs’ underlying medical conditions were unavailable. Additional investigations are warranted in larger populations with adjustment for underlying comorbidities, other factors predisposing to complications and clinical management. Second, although our results implied headache may be associated with more favorable outcomes, we cannot exclude the possibility that headache is more likely to be perceived by patients with mild illness.

Dr. Lan is a PhD candidate in Population Health Sciences at Harvard University, USA. He is also an occupational physician at National Cheng Kung University Hospital, Taiwan. His primary research interest involves environmental and occupational medicine and epidemiology.

## Data Availability

De-identified data are available upon request.

## Acknowledgments

All authors declare no competing interest, and none of the authors receives funding toward the present study.

## Notes

### Competing Interest Statement

The authors have declared no competing interest.

### Author Declarations

The Institutional Review Board of Cambridge Health Alliance reviewed the parent study protocol, determined it to be exempt and waived informed consent based on the use of existing, HIPAA-deidentified data.

